# Focal transcranial direct current stimulation of auditory cortex in chronic tinnitus: A randomized controlled mechanistic trial

**DOI:** 10.1101/2023.07.12.23292557

**Authors:** Amber M. Leaver, Yufen J. Chen, Todd B. Parrish

## Abstract

**Objective:** The goal of this pilot MRI study was to understand how focal transcranial direct current stimulation (tDCS) targeting auditory cortex changes brain function in chronic tinnitus.

**Methods:** People with chronic tinnitus were randomized to active or sham tDCS on five consecutive days in this pilot mechanistic trial (n=10/group). Focal 4×1 tDCS (central anode, surround cathodes) targeted left auditory cortex, with single-blind 2mA current during twenty-minute sessions. Arterial spin-labeled and blood oxygenation level dependent MRI occurred immediately before and after the first tDCS session, and tinnitus symptoms were measured starting one week before the first tDCS session and through four weeks after the final session.

**Results:** Acute increases in cerebral blood flow and functional connectivity were noted in auditory cortex after the first active tDCS session. Reduced tinnitus loudness ratings after the final tDCS session correlated with acute change in functional connectivity between an auditory network and mediodorsal thalamus and prefrontal cortex. Reduced tinnitus intrusiveness also correlated with acute change in connectivity between precuneus and an auditory network.

**Conclusions:** Focal auditory-cortex tDCS can influence function in thalamus, auditory, and prefrontal cortex, which may associate with improved tinnitus.

**Significance:** With future refinement, noninvasive brain stimulation targeting auditory cortex could become a viable intervention for tinnitus.

**HIGHLIGHTS:**

- Focal transcranial direct current stimulation (tDCS) of auditory cortex changes cerebral blood flow and connectivity in tinnitus
- Tinnitus loudness ratings decreased on average after five sessions of active focal tDCS
- Acute changes in auditory, thalamic, and prefrontal function may predict quieter tinnitus after five sessions

## INTRODUCTION

Chronic tinnitus is a common condition, where people perceive a constant ringing or buzzing sound in the absence of a physical source for that percept (Heller, 2003; Jarach et al., 2022; McCormack et al., 2016). Acute, temporary tinnitus is common (e.g., after acoustic trauma, brain injury, or stress), and some forms of tinnitus have a known medical origin like acoustic neuroma or vascular malformation that can be treated. However, some patients develop chronic tinnitus with no clear cause, for which very few effective treatments are available. This type of chronic subjective tinnitus can significantly impact quality of life, and can sometimes lead to disability, mental health issues, and suicidality (Han et al., 2018). Many treatments have been explored, e.g., hearing aids, acoustic/noise therapy, medications, or cognitive behavioral therapy, some of which provide distraction and/or improved quality of life (Baldo et al., 2012; Cima et al., 2012; Hesser et al., 2011; Hoare et al., 2014). However, no currently available treatments consistently and safely reduce tinnitus loudness or intrusiveness (Langguth et al., 2019), and new effective treatments are needed.

It is unclear why some patients develop chronic tinnitus while others do not. Animal studies show hyperactivity and increased synchrony within the ascending auditory pathway in animal models of tinnitus (Basura et al., 2015; Eggermont and Roberts, 2004; Engineer et al., 2011; Kaltenbach et al., 2004; Shore et al., 2016); these results have been corroborated by human neuroimaging studies (Gu et al., 2010; Leaver et al., 2011; Lockwood et al., 1998; Maudoux et al., 2012; Sedley et al., 2015). Human tinnitus research has also implicated brain regions and networks outside the auditory system, including frontal cortex (Leaver et al., 2012; Schlee et al., 2009; Seydell-Greenwald et al., 2014, 2012), thalamic nuclei (Leaver et al., 2016; Mühlau et al., 2006), parahippocampal cortex (Landgrebe et al., 2009; Vanneste et al., 2011), basal ganglia (Cheung and Larson, 2010; Hinkley et al., 2022; Leaver et al., 2016, 2011; Maudoux et al., 2012), and others. This has led some researchers to propose that these nonauditory regions/networks may be critical to tinnitus pathophysiology, perhaps reflecting the inability of these regions to regulate or suppress aberrant activity within the auditory system (e.g., frontal attention networks (Roberts et al., 2013), fronto-striatal evaluative networks (Leaver et al., 2011), and/or affect regulation networks (Jastreboff, 1990; Møller, 2003; De Ridder et al., 2011)). In sum, available research suggests that chronic tinnitus may be a problem of the brain and brain networks, not (only) of the ear.

Correspondingly, there is interest in improving symptoms by “correcting” these tinnitus-related differences with noninvasive brain stimulation (NIBS), many of which target auditory cortex (Kok et al., 2021; Liang et al., 2020; Schoisswohl et al., 2019; Soleimani et al., 2016). NIBS methods are noninvasive, avoid the systemic side effects of drugs, and have little-to-no long-term side effects beyond discomfort during the stimulation session (Bikson et al., 2016). One popular type of NIBS, transcranial magnetic stimulation (TMS), has been successful in improving tinnitus in some randomized controlled trials targeting auditory cortex (Folmer et al., 2015; Forogh et al., 2016; Marcondes et al., 2010), though results are mixed (Kleinjung et al., 2007; Landgrebe et al., 2017). Transcranial direct current stimulation (tDCS) is another type of NIBS that uses less stimulation than TMS, but is cheaper, less uncomfortable, and potentially portable. Some previous randomized controlled tDCS trials targeting auditory cortex also reported reduced tinnitus impact or loudness (Henin et al., 2016; Hyvärinen et al., 2016; Shekhawat et al., 2016); however, negative trials exist (Forogh et al., 2016), and considerable heterogeneity in protocol design (e.g., stimulation target(s), electrode shape and size, number of sessions) make interpreting this literature challenging (Martins et al., 2022). So although previous controlled auditory-cortex tDCS trials are promising, additional work is needed.

Understanding how tDCS changes brain activity could motivate improvements in future tDCS trials. Presumably, TMS or tDCS targeting auditory cortex could reduce tinnitus by decreasing aberrant auditory cortex activity, and/or by changing connectivity between auditory cortex and other parts of the brain like prefrontal cortex. Our own work shows that conventional, sponge-electrode tDCS targeting auditory cortex (left temporoparietal area) can reduce functional connectivity between prefrontal cortex and an auditory resting-state network during stimulation (Leaver et al., 2022). However, it is unclear how acute effects of tDCS during and immediately after one stimulation session relate to long-term changes in brain activity and/or symptoms after several sessions typically needed to improve symptoms. Studies combining neuroimaging with randomized-controlled application of tDCS and other NIBS methods could be illuminating.

In this mechanistic pilot trial, we used MRI to understand how focal tDCS targeting auditory cortex changes brain function in people with chronic tinnitus, particularly in cases where tinnitus improves after multiple stimulation sessions (**Figure 1**). We used 4×1 focal tDCS (center anode) using an established head-landmark method (Langguth et al., 2006b) to deliver more focal stimulation to primary auditory cortex compared with conventional sponge tDCS electrodes. Assignment was randomized, parallel, and blinded, which is particularly important given the subjective nature of tinnitus and the robustness of the placebo effect. Brain function was measured immediately before and after the first stimulation session using two complementary methods: blood oxygenation level dependent (BOLD) fMRI to measure functional connectivity, and arterial spin-labelled (ASL) MRI to obtain an absolute quantification of cerebral blood flow (CBF in 100g/mL/min). Changes in tinnitus loudness, intrusiveness, and overall impact (tinnitus functional index, TFI) were also measured after five tDCS sessions. We hypothesized that auditory-cortex tDCS would change CBF and functional connectivity within and between brain regions previously implicated in tinnitus, specifically auditory and prefrontal cortex (primary outcome). Though five sessions is far fewer than is typically used in efficacy trials of tDCS and TMS (e.g., in depression or obsessive-compulsive disorder (Brunoni et al., 2017; Carmi et al., 2019; George et al., 2010; Loo et al., 2018; O’Reardon et al., 2007; Zangen et al., 2021)), we also hypothesized that tinnitus would improve after active tDCS in some volunteers, and that the magnitude of these improvements would correlate with changes in brain function (secondary outcomes).

**Figure 1.**
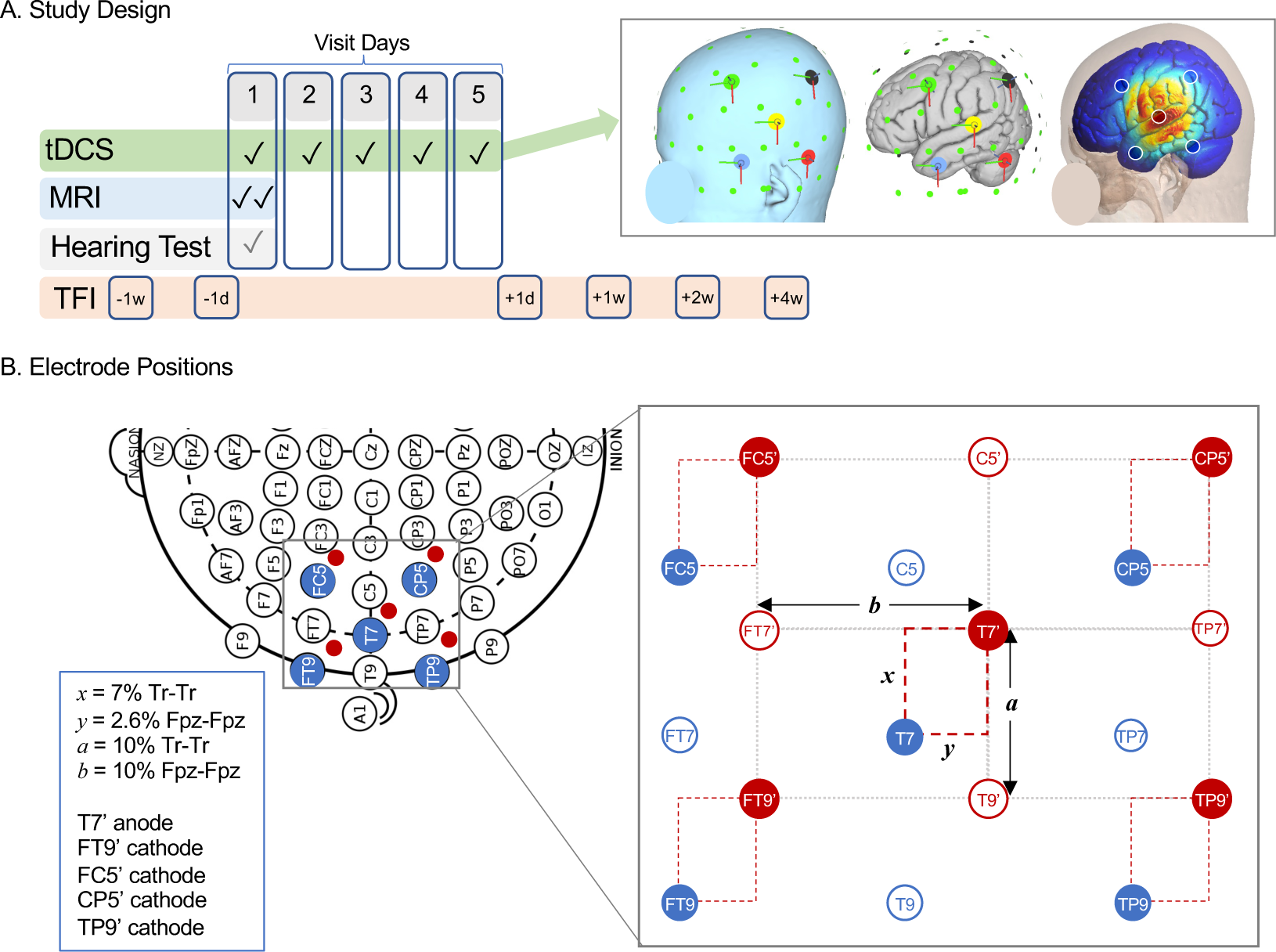
Study methods. **A.** Relative timing of tDCS sessions and key study assessments, including MRI, hearing test, and Tinnitus Functional Index (TFI) questionnaire are displayed. **B.** Schematic of electrode positioning method to target left primary auditory cortex (Heschl’s gyrus) is displayed on 10-10 EEG grid (left), with relevant standard positions marked in blue. Suggested stimulation site from Langguth et al. is 2.5cm superior and 1.5 posterior to position T7, which corresponds to distances *x* and *y* in the grid at right. Our study adjusted *x*and *y* to each volunteer’s head measurements, summarized at far right and Methods. Distance between anode (T7’) and cathodes (FT9’, FC5’, CP5’, and TP9’) was based on distance between T7 and T9 vertically (*a*) and FT9 horizontally (*b*).

## MATERIALS AND METHODS

### Subjects

This study was carried out in accordance with the Declaration of Helsinki after review and ethical approval of the Northwestern University Institutional Review Board, and all participants gave informed written consent before enrollment. Volunteers were recruited from the Chicago area using website advertisements and patient referrals. Enrollment began January 2022, and study ended as planned in July 2022. All study procedures took place at Northwestern University and Northwestern Medicine. Target sample size was n=20 for this mechanistic pilot trial; intention-to-treat sample achieved this goal (refer to CONSORT diagram and checklist in **Supplement**).

Inclusion/exclusion criteria were as follows. All participants were between 18 and 75 years old, and had chronic tinnitus for at least one year. Tinnitus must have been present (perceived) when consciously attended to for at least 50% of awake time, and intruded at least 10% of awake time, over the 12 months before screening. Participants must have reported discussing tinnitus symptoms with a physician or audiologist to rule out obvious physical or neurological origin for tinnitus (e.g., acoustic neuroma, Meniere’s Disease, vascular pulsatile tinnitus). If applicable, pharmacological treatment for tinnitus must not have changed 6 weeks prior to study start. Standard safety considerations for MRI and tDCS were exclusionary, including the presence of incompatible or unspecified implants, significant head injury, claustrophobia, scalp injury or other skin conditions, concurrent use of decongestants, antihistamines, benzodiazepines or other anticonvulsants, and anti-psychotics. Significant or severe developmental, neurological, or psychiatric conditions or substance abuse were also exclusionary, but mild-to-moderate mood or anxiety disorder or symptoms were not exclusionary.

### Trial Design

Summary of study schedule is displayed in **Figure 1A**, and study was registered at clinicaltrials.gov as NCT05120037. Participants were randomly assigned to receive active or sham stimulation using a random number generator in Microsoft Excel, stratified by sex and age (4 groups: 18-29, 30-44, 45-64, 65+ years) by A.M.L. Assignment was parallel. TDCS sessions were scheduled on five consecutive days, and MRIs were done before and after the first tDCS session. Tinnitus assessments were completed 1 week and 1 day before the first tDCS session (−1w, −1d), and 1 day, 1 week, 2 weeks, and 4 weeks after the final tDCS session (+1d, +1w, +2w, +4w). Volunteers rated tDCS-related paresthesia after each session, and common potential side effects were queried after tinnitus assessments (i.e., −1d, +1d, +1w, +2w, +4w).

Participants remained blind to group assignment until four weeks after the final tDCS session (i.e., +4w, after final round of surveys was complete). To preserve blinding, all volunteers received the same description of tDCS side effects before and during the study, and a privacy screen was placed between the volunteer and the tDCS device. In addition, all tinnitus assessments were completed using an online system (REDCap) by the participant after study visits on the participant’s personal device (i.e., not in the presence of the study team). One volunteer completed assessments on paper at home (−1w) and at NU (Visits 1 and 5 for −1d and +1d, respectively), and to a blinded study coordinator via phone interview (+1w, +2w, +4w). Blinding was assessed in the +4w follow-up in a survey including a binary guess (active or sham) and 4-point confidence scale (range: “very confident” to “not at all confident”).

MRI data were pre-specified primary outcomes, including change in functional connectivity and cerebral blood flow in auditory cortex. Secondary outcomes were changes in TFI score and their correlation with MRI data, including overall score, Tinnitus Awareness (TFI question #1) and Tinnitus Loudness (TFI question #2). Sample size was determined based on typical size of pilot MRI studies and available funding.

### TDCS Sessions

Focal tDCS was delivered using a Soterix 1×1 tES device with a 4×1 HD-tDCS conversion box. Electrodes were Ag/AgCl rings placed in custom plastic holders affixed to a spandex cap and filled with Lectron II conductive gel (∼1.5mL/electrode). Current amplitude was 2mA (i.e., +2mA anode, and −0.5mA each cathode), delivered with 30 second linear on- and off-ramps per manufacturer settings. Sessions were 20 minutes in duration; current was constant for active tDCS, and current was ramped up and down at the beginning and end of the 20-minute session for sham stimulation (as programmed by the manufacturer). Paresthesia (e.g., tingling, itching) tends to be greatest during the beginning of the session and during changes in current; transient stimulation in sham programming attempts to equate paresthesia for sham and active conditions in this regard.

Electrode configuration was 4×1, with center anode positioned over auditory cortex surrounded by four cathodes (**Figure 1**). Anode position followed the method developed by Langguth et al..(Langguth et al., 2006b) In brief, this method recommends stimulation at 2.5 cm superior and 1.5 cm posterior to position T7/T3 along the Tr-Cz and Fpz-Oz axes, respectively, within the 10-10/10-20 EEG system. In their analysis, this scalp position had the shortest Euclidean distance to Heschl’s gyrus in a group of 25 control volunteers measured on anatomical MRI. Because this method may be inaccurate for some head shapes/sizes (Noh et al., 2017; Theodoroff et al., 2018), we made small adjustments to this approach to accommodate head size and shape. Anode was positioned 7% of Tr-Tr distance superior and 2.6% of Fpz-Fpz distance posterior to T7. This yielded average distances similar to Langguth et al.; mean (SD) was 2.65 (0.12) and 1.48 (0.06) cm, respectively. Cathodes were placed around the anode in a rectangle with sides parallel to Fpz-Oz and Tr-Cz axes; distance from anode was based on T7-FT7 distance horizontally (mean(SD) = 5.68(0.21) cm) and T7-T9 distance vertically (mean(SD) = 3.78(0.18) cm).

### Clinical & Behavioral Assessments

Each volunteer underwent pure-tone threshold testing at Northwestern Medicine’s Audiology clinic, including standard frequencies and above 8kHz to capture potential high frequency hearing loss. Tinnitus assessments were done using self-report surveys, including Tinnitus Functional Index (TFI), Tinnitus Handicap Inventory (THI), and Tinnitus Sample Case History Questionnaire. Additional surveys included Beck Depression and Anxiety questionnaires, and basic demographic and medical history items relevant to the study. Changes in TFI scores were pre-selected secondary outcomes for this study, including total score (score range 0-100), TFI item 2 (loudness rating, 10 point scale) and TFI item 1 (% awake time aware, 10 point scale). Potential side effects of tDCS were measured using self-report questionnaires. Paresthesia during stimulation was rated after each visit on 10-point scales for intensity and discomfort. Headaches, fatigue, difficulty concentrating and sleep difficulties were assessed weekly beginning with the first tDCS session through 4-week follow-up.

### MRI Acquisition & Preprocessing

MR images were acquired using a 3T Prisma scanner and 64-channel head coil (Siemens) at the Northwestern University Center for Translational Imaging. Sequence parameters for BOLD-fMRI scans were as follows: 2mm^3^ isotropic resolution, 0.8s repetition time (TR), 37ms echo time (TE), 52° flip angle, 72 axial slices, anterior/posterior phase encoding, 8 multiband factor, 600 volumes (8.17s acquisition time). 3D GRASE, background-suppressed pseudo-continuous ASL images(Kilroy et al., 2014) were also acquired: 2.5×2.5×3mm^3^ resolution, TR 4.3s, TE 36.48ms, 1500ms label duration, 1800ms post-label delay, anterior/posterior phase encoding, 7 label/control volume pairs. A nonlabelled calibration M0 image was acquired immediately before the ASL volume pairs. T1- and T2-weighted anatomical scans were also acquired: T1 multi-echo MPRAGE 0.8mm isotropic, TR=2.17s, TE=1.8, 3.6, 5.4ms, 1160ms inversion time; 7° flip angle; T2 0.8mm isotropic, TR=3.2s, TE=564ms, echo train length = 1166ms. All raw and preprocessed images passed visual inspection for quality.

BOLD preprocessing was implemented using FSL software, including deletion of first 4 volumes, motion correction, high-pass temporal filtering (100s), and manual ICA-based denoising (rated by A.M.L.) (Griffanti et al., 2017; Salimi-Khorshidi et al., 2014). Spatial image registration used FSL’s Boundary-Based Registration (BBR), including correction for phase-encode direction and nonlinear registration to a standard MNI template (Greve and Fischl, 2009), followed by spatial smoothing at 4mm^3^. FSL’s dual regression procedure (Nickerson et al., 2017) calculated the strength of temporal coherence (functional connectivity) between resting brain activity (BOLD-fMRI timecourse) of each voxel and the timecourse of resting state networks (RSNs) defined using the Yeo Atlas (17 networks liberal mask (Thomas Yeo et al., 2011)). Finally, connectivity values for each RSN were averaged within each region using atlases described further below in volume space to improve signal-to-noise and reduce computational burden.

ASL preprocessing used FSL’s BASIL tool (Chappell et al., 2009). Images were corrected for motion using the calibration as a reference, and cerebral blood flow (CBF) was calculated using standard parameters (Alsop et al., 2015) and with correction for voxelwise M0 values derived from the calibration image. Spatial image registration used FSL’s BBR, including nonlinear registration to a standard MNI template and spatial smoothing at 4mm^3^. Finally, CBF was averaged within each region using the same atlases for BOLD data described further below.

### Statistical Analyses

All statistical analyses were completed in R (https://www.r-project.org), including the lme4 package (Bates, Douglas et al., 2015). Age, sex, and mean pure-tone audiometric threshold did not vary between groups (**Table 1**) or over time; therefore, these were not used as nuisance factors in statistical models. Statistical tests of relevant demographic and clinical variables were done using t or Chi-square tests to test for differences between active and sham groups.

**Table 1.**
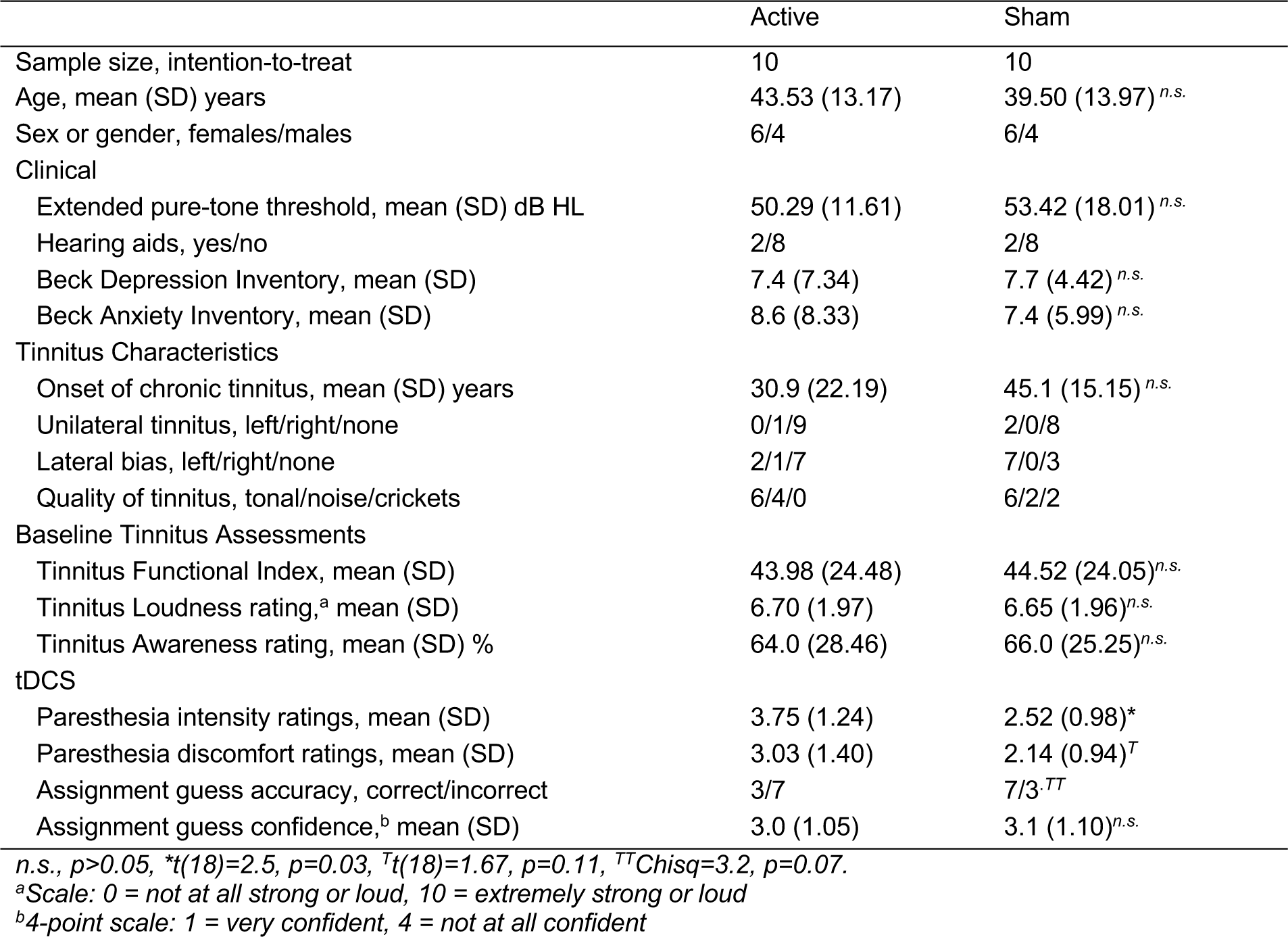
Demographic, Clinical, and tDCS Information

Change in tinnitus symptoms were tested using a time-by-group interaction in linear mixed models (Bates, Douglas et al., 2015), with time as a numerical factor, group as a categorical factor, and subject as random factor. Tinnitus symptoms included total TFI score, Tinnitus Loudness (self-reported loudness on 10-point scale; TFI question #2), and Tinnitus Awareness (% time aware of tinnitus on 10-point scale; TFI question #1). Baseline scores were defined as the average of ratings given 1 week before and 1 day before the first tDCS session. If a time-by-group interaction was not present at p<0.05, we also tested for a main effect of time. In cases of interaction or main effect of time p<0.05, change over time was assessed pairwise over time in each group separately (Tukey HSD correction for p values).

For MRI statistics, ASL and BOLD metrics were tested iteratively across atlas-defined regions, including Schaeffer et al. 400 cortical atlas (Schaefer et al., 2018), Behrens et al. thalamic atlas (Behrens et al., 2003), Choi et al. basal ganglia atlas (Choi et al., 2012), Buckner et al. cerebellar atlas (Buckner et al., 2011), and FSL’s Harvard-Oxford subcortical atlas for remaining structures. Within each of these regions, we calculated mean CBF for ASL MRI and mean functional connectivity between each region and each of 17 RSNs defined by Yeo et al. (Thomas Yeo et al., 2011) for BOLD fMRI. Two statistical models were used. The first planned analysis used linear mixed models to identify changes in brain function after active tDCS vs. after sham tDCS using a group-by-time interaction. Here, time was before/after the first tDCS session, group was a categorical factor (active or sham), and subject was a random factor. ASL and BOLD metrics were dependent variables. In cases where an interaction met criteria for significance (described further below), post-hoc pairwise comparisons tested for change over time separately for each group, uncorrected p<0.05.

The second planned MRI analysis used linear models to identify changes in brain function after active tDCS that correlated with change in tinnitus symptoms after five active tDCS sessions. Here, the dependent variable was change in tinnitus symptom score. We selected changes in tinnitus symptoms that met statistical criteria described above for tinnitus assessments (i.e., time-by-group interaction p<0.05 and/or post-hoc pairwise difference over time, Tukey-HSD-p<0.05). Linear models targeted an interaction between change in MRI metric (before/after the first tDCS session) and group (active or sham), both fixed factors. For each result meeting criteria for significance (described further below), a post-hoc Pearson correlation assessed the relationship between change in tinnitus symptom and change in MRI metric separately for each group (active or sham), uncorrected p<0.05.

Criteria for statistical significance were more strict for exploratory analyses than for planned analyses. Change in auditory cortex function after active tDCS was a pre-specified primary outcome for this trial. Therefore, we report any interaction present in auditory cortex p_uncorr_ <0.05 for CBF and functional connectivity (FC) with the auditory RSN. We also report any interaction p_uncorr_ < 0.005 between the auditory RSN and any brain region. Early auditory regions are displayed in **Figure 3**. In exploratory analyses of every brain region and network, results were corrected for false discovery rate q<0.05 across regions within each MRI metric (CBF, each RSN).

## RESULTS

### Behavioral & Clinical outcomes

Volunteers assigned to active or sham tDCS groups did not differ with respect to age, sex, mean pure-tone audiometric threshold, hearing aid use, mood disorder symptoms, age of tinnitus onset, or baseline tinnitus symptom severity (i.e., TFI score, Tinnitus Loudness rating, and Tinnitus Awareness rating; **Table 1**). Mean ratings of tDCS-related paresthesia intensity were higher after active tDCS, and discomfort ratings were also marginally higher after active tDCS. Volunteers in the active and sham groups guessed “sham” as their group assignment with equal frequency, making the active tDCS group marginally less accurate in guessing their group assignment four weeks after the final tDCS session. Confidence ratings for group assignment guesses did not differ between groups (**Table 1**). Intensity ratings for headaches, fatigue, sleep difficulty, and concentration difficulty did not differ at baseline between groups, and did not change differently over time (**Supplemental Table 1**).

Change in tinnitus assessments over time did not differ between active and sham groups in statistical models including all baseline and follow-up timepoints (i.e., all time-by-group interactions p > 0.05). However, all three assessments showed main effects of time (**Table 2**, **Figure 2**). In pairwise comparisons over time in each group separately, Tinnitus Loudness ratings were significantly lower two weeks after active tDCS compared with baseline scores (Cohen’s d (SE) = 1.49(0.46)) and compared with scores immediately after the final tDCS session (Cohen’s d (SE) = 1.58(0.46)). Tinnitus Awareness was also significantly lower four weeks after active tDCS compared with baseline (Cohen’s d (SE) = 1.47(0.46)). These significant pairwise differences in Tinnitus Loudness and Tinnitus Awareness after active tDCS were used in MRI correlations described further below. For Tinnitus Loudness, the difference with larger effect size was used in MRI analyses (i.e., difference in loudness rating between +0d and +2w). All descriptive and pairwise statistics are given in **Supplemental Tables 1 and 2**, respectively.

**Figure 2.**
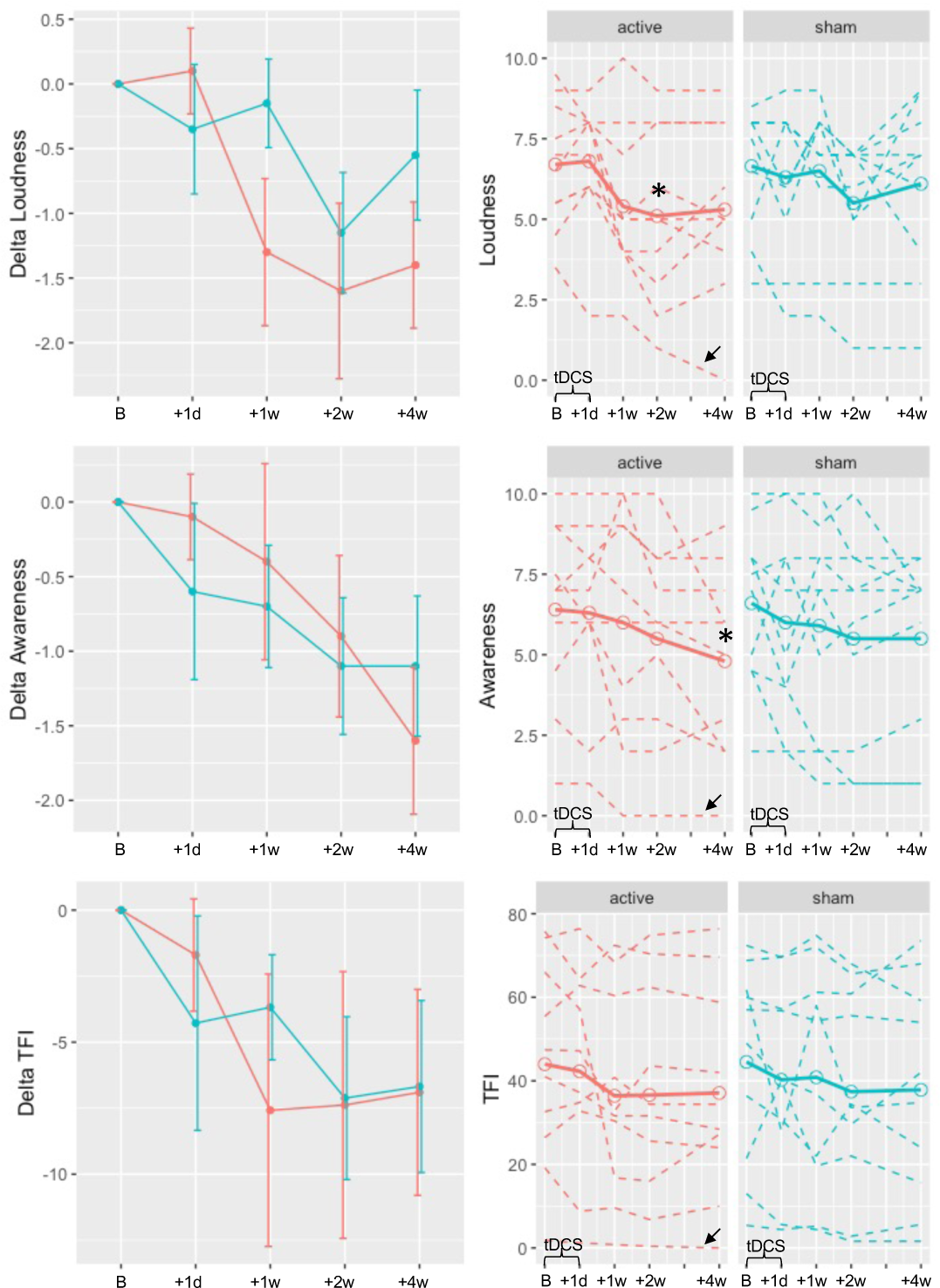
Changes in tinnitus scores after active and sham tDCS. Tinnitus scores are plotted for Tinnitus Functional Index (TFI) Loudness rating (item #2, top), TFI Tinnitus Awareness rating (item #1, middle), and overall score (bottom). Time is plotted on x axes in weeks relative to first tDCS session, with time B representing baseline average over assessments −1w and −1d, and corresponding to assessments +1d, +1w, +2w, and +4w as in Figure 1A. In left column, mean change in rating score is plotted for active (red) and sham (blue); error bars are standard error. In right column, mean rating is plotted in open circles and solid lines, and each volunteer’s ratings are plotted in dashed lines. Asterisks mark times significantly different from baseline (p-corr < 0.05). Outlier with low baseline TFI and enlarged ventricles marked by black arrow.

**Table 2.** Change in Tinnitus Assessments

| Assessment | Statistical Test | Full Sample, n=10/group |  |  |  |  | Omitting outlier, 9 active, 10 sham |  |  |  |  |
| --- | --- | --- | --- | --- | --- | --- | --- | --- | --- | --- | --- |
| | | $\beta$ | $\beta$ SE | $t$ | $p$ | $p_{Tukey}$ | $\beta$ | $\beta$ SE | $t$ | $p$ | $p_{Tukey}$ |
| TFI | time*group | 0.21 | 0.87 | 0.25 | 0.81 | N/A | 0.34 | 0.91 | 0.37 | 0.71 | N/A |
|  | time | -1.46 | 0.61 | -2.39 | 0.02 | N/A | -1.59 | 0.66 | -2.40 | 0.02 | N/A |
| Loudness | time*group | 0.21 | 0.13 | 1.58 | 0.12 | N/A | 0.17 | 0.14 | 1.26 | 0.21 | N/A |
|  | time | -0.34 | 0.09 | -3.71 | 0.0004 | N/A | -0.31 | 0.10 | -3.11 | 0.003 | N/A |
|  | active, baseline vs. +2wk | 1.60 | 0.48 | 3.33 | 0.001 | 0.04 | 1.50 | 0.51 | 2.94 | 0.004 | 0.11 |
|  | active, +0d vs. +2wk | 1.70 | 0.48 | 3.53 | 0.0007 | 0.02 | 1.78 | 0.51 | 3.48 | 0.0009 | 0.03 |
| Awareness | time*group | 0.13 | 0.12 | 1.04 | 0.30 | N/A | 0.14 | 0.13 | 1.09 | 0.28 | N/A |
|  | time | -0.34 | 0.09 | -3.89 | 0.0002 | N/A | -0.35 | 0.09 | -3.73 | 0.0004 | N/A |
|  | active, baseline vs. +4wk | 1.60 | 0.49 | 3.29 | 0.002 | 0.05 | 1.67 | 0.52 | 3.18 | 0.002 | 0.06 |

During analysis, a potential outlier was identified in the active group, with low baseline TFI score and enlarged ventricles. Omitting this volunteer did not have a large impact on statistics for tinnitus assessments (**Table 2**). However, we report MRI analyses omitting this outlier due to concerns regarding spatial normalization of MRI data.

### MRI outcomes

For planned analyses in early auditory regions, significant time-by-group interactions were present in left Heschl’s gyrus for CBF and bilaterally for FC with the auditory resting state network (AUN) (p<0.05; **Figure 3**, **Table 3**). In post hoc tests, AUN FC and CBF increased after active tDCS in these regions, but did not change after sham tDCS. Descriptive and pairwise statistics are given in **Supplemental Tables 3 and 4**, respectively. A time-by-group interaction was also noted between AUN and a region in right dorsal lateral occipital cortex (p<0.005), which appeared to be driven by increased FC in the sham group. Time-by-group interactions were not present for CBF or FC in any other regions or networks at our chosen threshold criteria.

**Figure 3.**
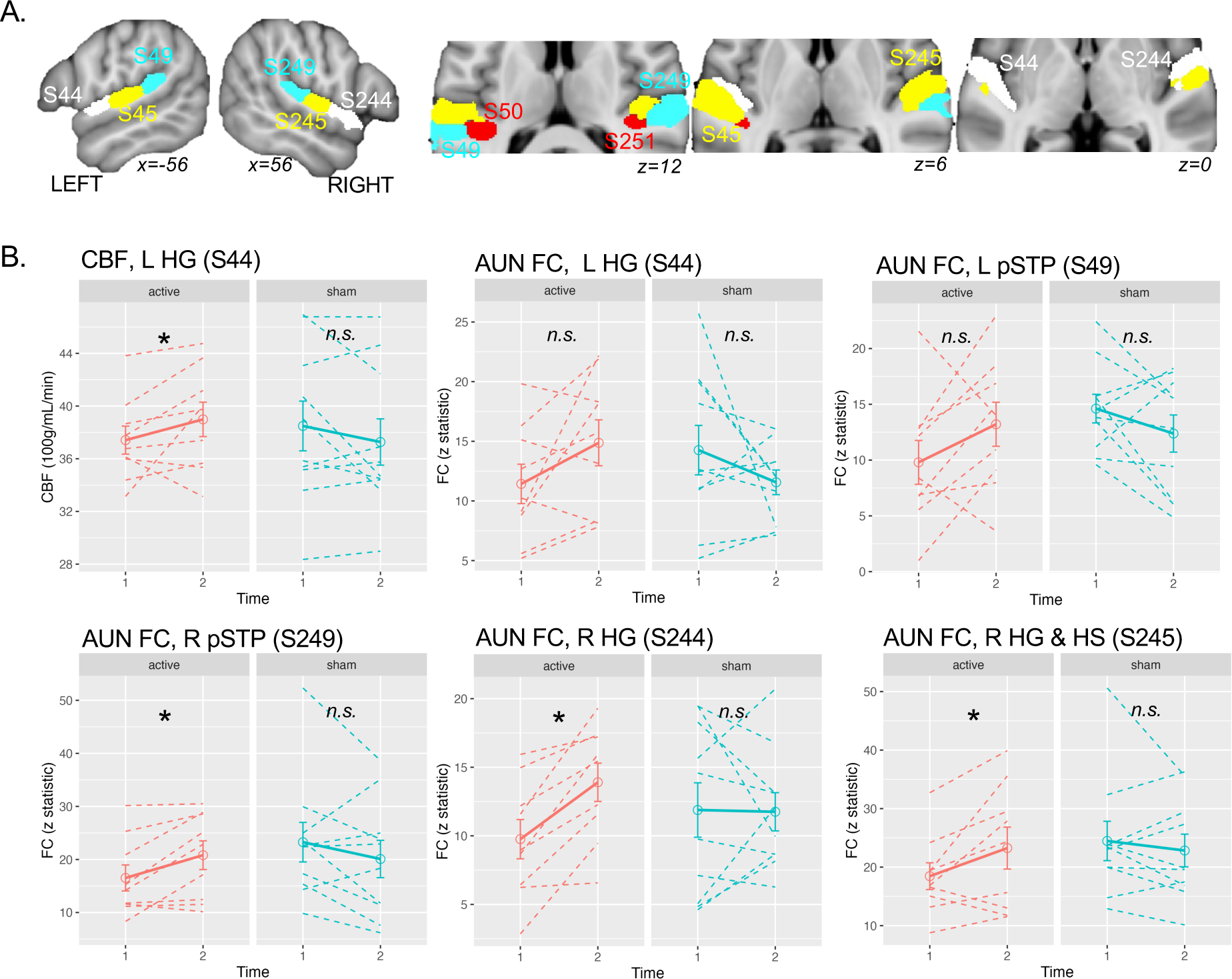
Acute increases in brain function in auditory cortex after the first active tDCS session. **A.** Early auditory cortical regions from the Schaeffer400 atlas (S) are displayed. **B.** Lineplots are displayed for each significant time-by-group interaction effect. X axes display Time 1 (before first tDCS session) and Time 2 (after first session), separately for active (red) and sham (blue) groups. Solid lines are the group average, and single-subject data are plotted in dashed lines. Error bars are standard error. Asterisks mark post-hoc pairwise difference over time p < 0.05 (see detailed stats in Table 3). Abbreviations: R right, L left, medial, m; Heschl’s gyrus, HG; Heschl’s sulcus, HS; posterior superior temporal plane, pSTP; auditory network, AUN.

**Table 3.**
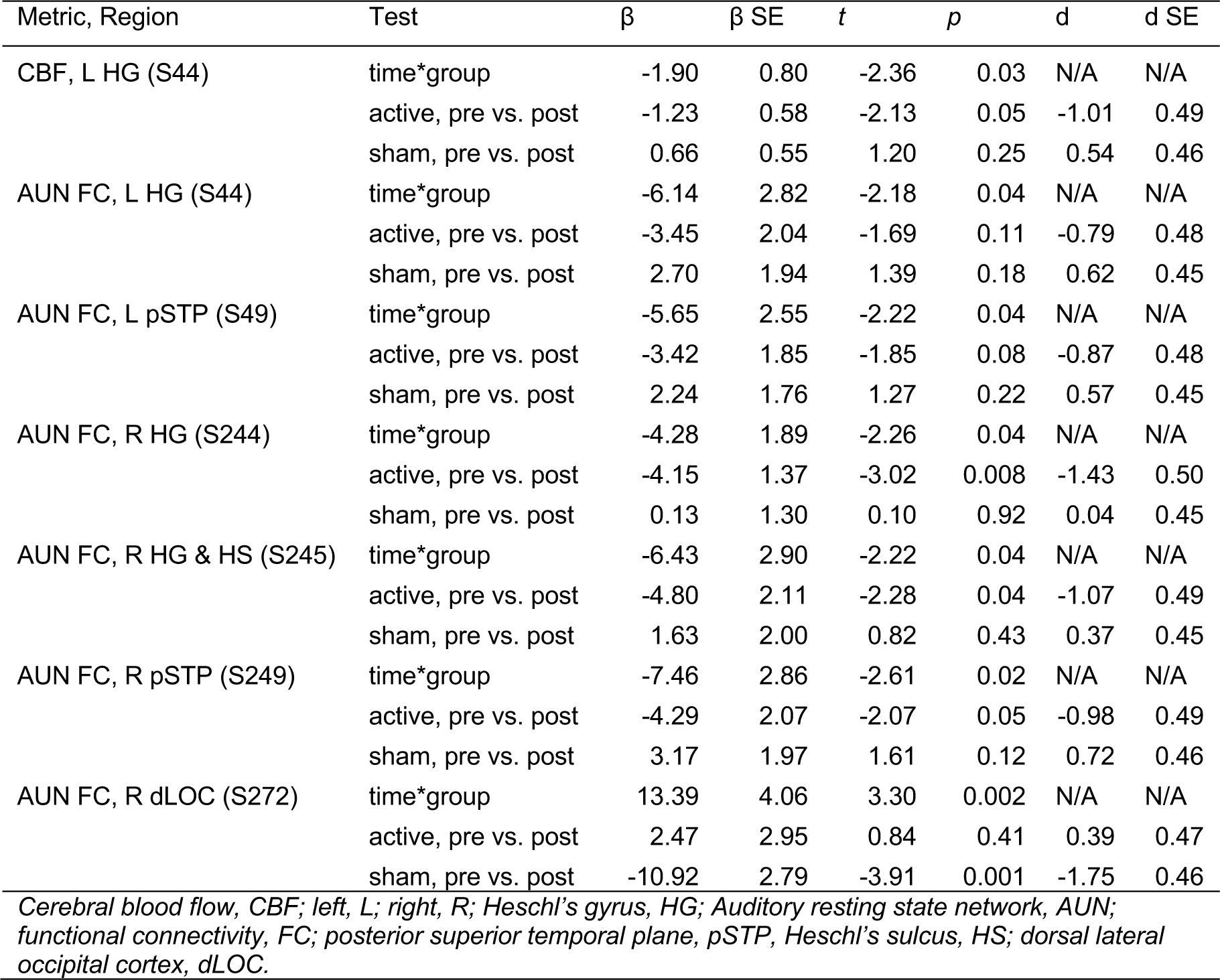
Acute change in MRI metrics after active vs. sham tDCS

Improved Tinnitus Loudness two weeks after the last active tDCS session (i.e., +0d vs. +2w) correlated with acute changes (Δ) in brain function after the first tDCS session (**Figure 4**, **Table 4**). In a planned analysis of AUN FC, a significant group-by-ΔFC interaction was noted for FC between AUN and right premotor cortex, dorsolateral prefrontal cortex, and bilateral thalamus “Sensory” division of the Behrens et al. atlas (p<0.005). Here, lower Tinnitus Loudness after active tDCS associated with increased AUN FC in the thalamus and decreased AUN FC in right prefrontal regions; modest opposing effects were noted for the sham group. These interactions were not present for any other regions or networks at our chosen criteria. However, subthreshold interactions were noted for CBF in the same “Sensory” division of the thalamus (p=0.008), as well as nearby “Primary Motor”, and “Pre-motor” divisions (p<0.005; **Supplemental Figure 3, Supplemental Table 5**). These thalamic atlas divisions overlap lateral medial nuclei (mediodorsal lateral parvocellular) and lateral nuclei (ventral posterolateral) when consulting histological/MRI atlases.(Behrens et al., 2003; Iglesias et al., 2018; Saranathan et al., 2021)

**Figure 4.**
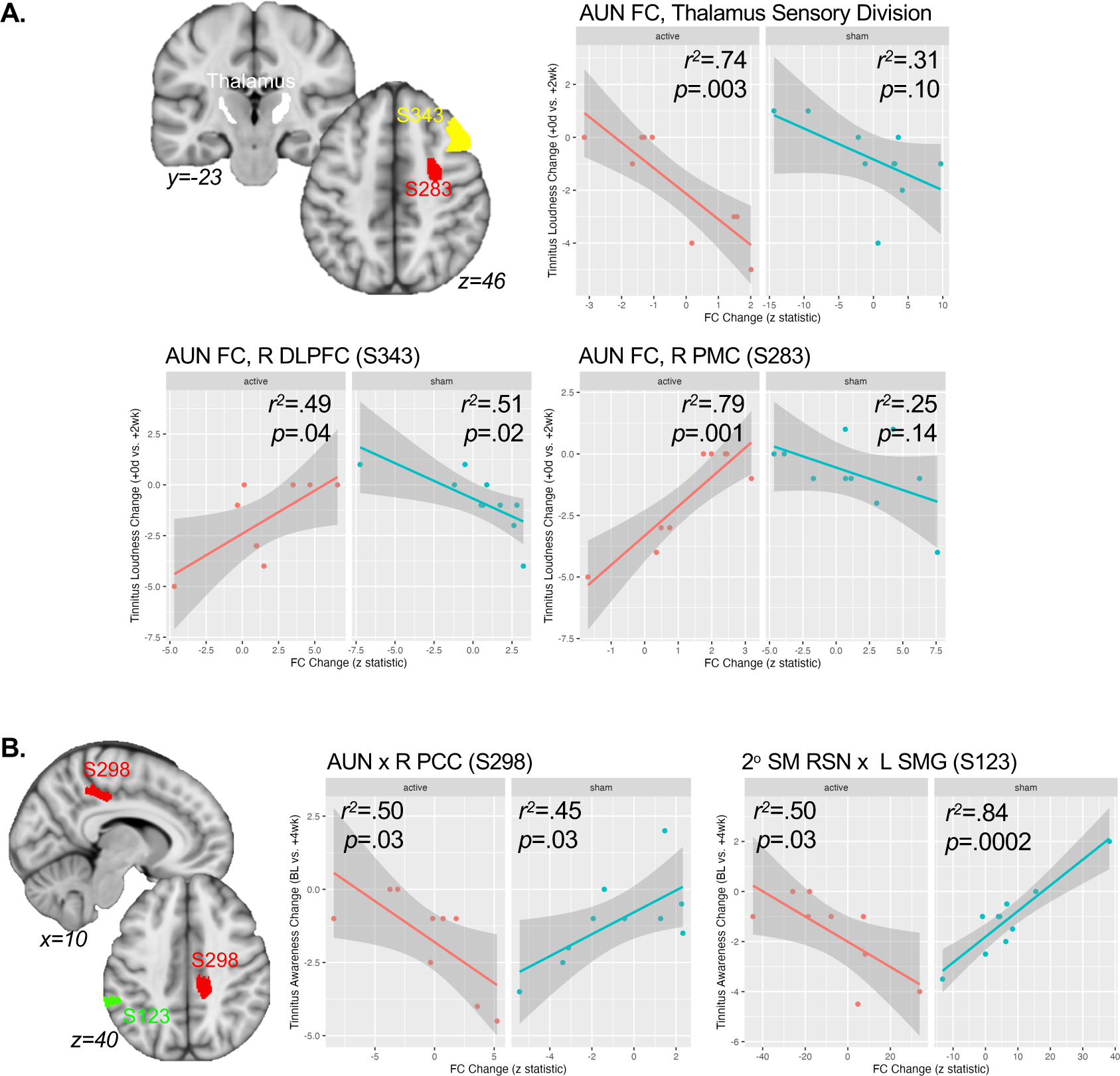
Acute changes in functional connectivity (FC) after the first active tDCS session correlate with reduced tinnitus after five sessions. **A.** Regions showing significant time-by-!Loudness interactions in FC with an auditory resting-state network (AUN). Scatterplots are displayed for each region, separately for active (red) and sham (blue) groups. **B.** Regions showing significant time-by-ΔAwareness interactions in FC. ΔAwareness is plotted using a 10-point scale (e.g., −2.5 is a 25% reduction in time spent aware of tinnitus). Scatter plots are shown as in A. Abbreviations: R right; dorsolateral prefrontal cortex, DLPFC; premotor cortex, PMC; auditory network, AUN; Schaeffer400 parcel, S; posterior cingulate cortex, PCC, secondary somatomotor resting state network, 2° SM RSN; supramarginal gyrus (SMG).

**Table 4.** Associations between acute change in MRI metrics and change in tinnitus assessments after active vs. sham tDCS

| Analysis | Metric, Region | Test | $\beta$ | $\beta$ SE | $t$ | $p$ | $r^2$ |
| --- | --- | --- | --- | --- | --- | --- | --- |
| ΔLoudness | AUN FC, R DLPFC (S283) | ΔFC*group | -1.37 | 0.30 | -4.57 | 0.0004 | N/A |
|  |  | active | 1.19 | 0.23 | 5.12 | 0.001 | 0.79 |
|  |  | sham | -0.18 | 0.11 | -1.65 | 0.14 | 0.25 |
| | AUN FC, R DLPFC (S343) | ΔFC*group | -0.77 | 0.20 | -3.79 | 0.002 | N/A |
|  |  | active | 0.42 | 0.17 | 2.54 | 0.04 | 0.48 |
|  |  | sham | -0.35 | 0.12 | -2.90 | 0.02 | 0.51 |
| | AUN FC, Thalamus (S436) | ΔFC*group | 0.85 | 0.25 | 3.43 | 0.004 | N/A |
|  |  | active | -0.97 | 0.22 | -4.46 | 0.003 | 0.74 |
|  |  | sham | -0.12 | 0.06 | -1.88 | 0.10 | 0.31 |
| ΔAwareness | AUN FC, R PCC (S298) | ΔFC*group | 0.65 | 0.18 | 3.59 | 0.003 | N/A |
|  |  | active | -0.28 | 0.11 | -2.64 | 0.03 | 0.50 |
|  |  | sham | 0.37 | 0.15 | 2.55 | 0.03 | 0.45 |
| | 2° SMN FC, L SMG (S123) | ΔFC*group | 0.15 | 0.03 | 5.42 | 0.0001 | N/A |
|  |  | active | -0.05 | 0.02 | -2.66 | 0.03 | 0.50 |
|  |  | sham | 0.10 | 0.02 | 6.52 | 0.0002 | 0.84 |

*Auditory resting state network, AUN; functional connectivity, FC; right, R; dorsolateral prefrontal cortex, DLPFC; posterior cingulate cortex, PCC; left, L; supramarginal gyrus, SMG.*

Reduced Tinnitus Awareness ratings four weeks after active tDCS (Baseline vs. +4w) also correlated with acute changes in brain function after the first tDCS session (**Figure 4**, **Table 4**). A significant group-by-ΔFC interaction was noted for FC between the AUN and right posterior cingulate cortex (PCC, p<0.005), where decreased ratings associated with increased connectivity after active tDCS. We also noted a group-by-ΔFC interaction for FC between a secondary somato-motor RSN and left supramarginal gyrus (p_fdr_<0.05), which was driven by a highly significant association in the sham group and only modest correlation in the active group.

## DISCUSSION

In this mechanistic pilot trial, we measured changes in MRI (CBF and FC) metrics and tinnitus ratings after focal tDCS targeting left auditory cortex. After the first active tDCS session, MRI metrics increased in early auditory cortex, including CBF in left Heschl’s gyrus and connectivity with an auditory resting state network (AUN) bilaterally. These findings are consistent with evidence that anodal electrical stimulation increases cortical excitability in targeted regions (Creutzfeldt et al., 1962; Purpura and McMurtry, 1965). Modest improvements in tinnitus metrics were also noted after five sessions of active tDCS, including lower tinnitus loudness ratings 2 weeks after the final stimulation session and decreased tinnitus awareness 4 weeks after the final session. Notably, acute changes in connectivity between AUN, thalamus, and prefrontal cortex after the first session correlated with (or predicted) these later improvements. Increased connectivity between AUN and PCC also correlated with later improvements in tinnitus intrusiveness (awareness). Taken together, these results suggest that anodal tDCS of auditory cortex acutely increases auditory cortical function in people with tinnitus, and downstream consequences of this in thalamus, prefrontal, and other regions may be important for later clinical response. Refining the application of tDCS to enhance these brain-network changes in all patients may improve the efficacy of tDCS to treat tinnitus in future studies.

### Auditory tDCS acutely increases auditory cortical function in tinnitus

Anodal tDCS is largely thought to increase cortical excitability under positive electrode(s) (Creutzfeldt et al., 1962; Purpura and McMurtry, 1965), an idea supported by previous work in the motor system (Nitsche and Paulus, 2001, 2000). In our study, both CBF and AUN FC increased in left early auditory cortical regions immediately after anodal tDCS targeting left auditory cortex. However, AUN FC also increased in early auditory regions in the right hemisphere, demonstrating that unilateral stimulation can influence auditory cortex function bilaterally. Effect sizes were also bigger in the right hemisphere, though we did not compare hemispheres quantitatively. This is consistent with the idea that unilateral stimulation of auditory cortex can influence both ipsalateral and contralateral activity; Matsushita et al. noted that unilateral auditory-cortex tDCS modulated bilateral sound-evoked EEG potentials (Matsushita et al., 2021), and Andoh, et al. demonstrated that a single session of continuous theta burst TMS targeting right auditory cortex increased sound-evoked activity and functional connectivity in contralateral auditory cortical regions (Andoh et al., 2015; Andoh and Zatorre, 2013). Though unilateral lesion or NIBS of somatosensory or motor cortex has clear, systematic effects on contralateral somatomotor function, no such clear relationship exists for human auditory cortex (Zatorre and Penhune, 2001). Therefore, it could be possible that bilateral NIBS is not necessary to induce bilateral effects in auditory cortical activity, and unilateral NIBS may have similar effects on tinnitus perceived in the right ear, left ear, or bilaterally. However, our study is not designed or powered to address these issues, and future, larger-scaled studies designed to answer these questions are needed.

Though details vary, overwhelming evidence supports auditory system involvement in chronic tinnitus, typically increased activity or function. In particular, molecular imaging studies reporting increased brain activity in left posterior auditory cortex (Arnold et al., 1996; Eichhammer et al., 2007; Langguth et al., 2006a; Song et al., 2012), motivated studies targeting auditory cortex with 1Hz rTMS (Langguth et al., 2006b), which is considered to be inhibitory. It may seem counterintuitive that anodal tDCS that increases (presumably) already hyperactive auditory cortex would lead to improved tinnitus symptoms in our study. However, it is possible that acute increases after the first tDCS session were followed by decreased auditory cortical function after several sessions not measured in this study, perhaps coinciding with improved symptoms weeks after treatment. It is also possible that these acute increases in auditory cortex are not directly related to later symptom improvements; indeed, they were not correlated with clinical outcomes in our study. Again, future studies that measure changes in brain function in the days and weeks after the end of treatment would be helpful in determining the timing of these acute and long-term changes in relation to improved tinnitus.

### Role of nonauditory region in response to noninvasive brain stimulation in tinnitus

The auditory system is clearly affected in tinnitus, but there is also evidence of functional and structural differences in people with tinnitus in other regions like prefrontal cortex (Schlee et al., 2009; Seydell-Greenwald et al., 2014, 2012), thalamus (Brinkmann et al., 2021; Leaver et al., 2016; Mühlau et al., 2006), parahippocampal cortex (Landgrebe et al., 2009; Vanneste et al., 2011), and basal ganglia (Cheung and Larson, 2010; Leaver et al., 2016, 2011; Maudoux et al., 2012). It is difficult to say whether these differences, including effects identified by our current study, are the cause or consequence of chronic tinnitus. In other words, these effects could reflect abnormalities that cause tinnitus to become (or remain) chronic, or they could reflect “normal” systems attempting to compensate for a chronic tinnitus caused elsewhere (e.g., within the auditory brainstem). Regardless, understanding how to modulate these networks to improve tinnitus symptoms could fill an important clinical need.

In our study, quieter tinnitus after five tDCS sessions appeared to be predicted by functional changes in thalamus and prefrontal cortex after the first tDCS session. Specifically, acute changes in connectivity between the auditory resting state network (AUN) and thalamus and right prefrontal regions correlated with reduced tinnitus loudness ratings after five sessions. Effect sizes were relatively high, with change in FC explaining 74% of variance in changed loudness rating for thalamus, and 79% of variance for right premotor cortex. Similar subthreshold effects were also present for CBF (Supplemental Figure 3), where quieter tinnitus associated with increased CBF in the same region of the thalamus near ventrolateral and mediodorsal subnuclei (MDN). Overall, these results are consistent with evidence that fronto-thalamo-striatal circuits function differently in chronic tinnitus, and that modulating these circuits may make tinnitus quieter. Indeed, combined stimulation of prefrontal and auditory regions with tDCS or rTMS has also improved tinnitus in some studies (Jacquemin et al., 2021, 2018; Noh et al., 2020), though further study is needed (Cardon et al., 2022).

Previous tinnitus neuroimaging studies have identified tinnitus-related differences in function and/or structure of thalamus, prefrontal cortex, and basal ganglia. In a previous fMRI study, we demonstrated modestly reduced connectivity between MDN and an auditory RSN, and identified a potential tinnitus network involving MDN, PFC, basal ganglia, and auditory cortex (Leaver et al., 2016). Notably, connectivity between MDN and this putative tinnitus network correlated with tinnitus loudness in this previous study (Leaver et al., 2016). The mediodorsal subnuclei of the thalamus primarily communicate with prefrontal cortex, and are thought to modulate cortico-cortical communication (Sherman and Guillery, 2013) to support various cognitive functions including associative learning, decision making, and others (Mitchell, 2015; Pergola et al., 2018). However, both ventrolateral and mediodorsal nuclei participate in thalamo-striato-frontal circuits that could be in a position to modulate auditory cortical activity. Ventrolateral nuclei communicate with motor and premotor cortex, and are involved in speech production (Hebb and Ojemann, 2013). Given that the motor system can modulate/suppress auditory-system activity during speech production (Paus et al., 1996), this circuit could also be relevant for tinnitus pathophysiology and/or treatment (and could also explain cases of somatic modulation of tinnitus through facial, head, or neck movement). It is difficult to delineate thalamic nuclei on standard MRI, so studies using specialized imaging may be needed to identify the precise thalamic subnuclei involved (e.g., submillimeter resolution (Saranathan et al., 2021)).

Decreased awareness of tinnitus was associated with increased connectivity between AUN and PCC in our study. The PCC is an important node in default mode networks, which have been implicated in awareness, self-referential processing, neuropsychiatric conditions like depression, among others (Raichle, 2015; Sheline et al., 2009). The PCC and adjacent medial parietal cortex in particular have been linked to attention and interoceptive awareness (Craig, 2003; Critchley et al., 2004), which our current results support. It is possible that the PCC and associated circuits mediate intrusiveness of the tinnitus into conscious perception. However, a modest opposing effect was also present in the sham group in our study (i.e., decreased awareness was associated with decreased connectivity between AUN and PCC), making this overall effect difficult to interpret. Validation of this and other study results with larger, independent studies is needed.

### Limitations and Conclusions

Overall, this first test of multi-session, focal tDCS targeting auditory cortex provides evidence that noninvasive brain stimulation of auditory cortex can modulate brain-network activity to improve the perceptual characteristics of tinnitus. Both tinnitus loudness and awareness were modestly reduced in the weeks following the final active tDCS session, consistent with extended response after multiple sessions of tDCS or TMS in some previous tinnitus studies (Bilici et al., 2015; Folmer et al., 2015; Forogh et al., 2016). Given that efficacy trials for tDCS and TMS in other conditions like depression typically include twenty or more sessions (Brunoni et al., 2017; Carmi et al., 2019; Cole et al., 2022; George et al., 2010; Loo et al., 2018; O’Reardon et al., 2007; Zangen et al., 2021), the stimulation “dose” used in our pilot study (five sessions) was likely inadequate to support robust, sustained improvements in tinnitus. Indeed, many tinnitus tDCS and TMS studies could be considered pilot studies in this regard, typically using 10 or fewer sessions (including single sessions), and it is possible that adding sessions to these protocols could lead to larger, more durable improvements. Perhaps more importantly, our study also provided clues as to why some volunteers improved after tDCS while others did not. In particular, modulating connectivity between the thalamus, prefrontal cortex and auditory network may be important in reducing perceived loudness of chronic tinnitus. Future, larger-scale studies may be needed to determine why these networks changed in some patients and not others; perhaps improved targeting of auditory cortex and/or other brain regions like DLPFC would improve engagement of these networks and subsequent clinical outcomes. There are limitations to our study that should be considered, including the tDCS protocol (e.g., limited number of sessions, head-landmark electrode placement, no personalization of stimulation amplitude), tinnitus assessments (e.g., no tinnitus loudness matching, minimum masking levels), and overall design (e.g., limited sample size, no long-term follow-up MRI). Given our modest sample size, we were also not able to measure potential effects of hearing loss severity, laterality or other qualities of the tinnitus percept, and other factors relevant to tinnitus (e.g., hyperacusis, somatic involvement, mood/anxiety disorder). Despite these limitations, our pilot study demonstrates the utility of longitudinal neuroimaging in furthering a mechanistic understanding of noninvasive brain stimulation in tinnitus and other conditions.

## Data Availability

All data produced in the present study are available upon reasonable request to the authors.

## ACKNOWLEDGEMENTS

This work was supported by the National Institutes of Health under award number DC015880 to Dr. Leaver. This content is solely the responsibility of the authors and does not necessarily reflect the official views of the National Institutes of Health.

## CONFLICT OF INTEREST

All authors declare no conflicts of interest.

## Notes

### Competing Interest Statement

The authors have declared no competing interest.

### Clinical Trial

NCT05120037

### Author Declarations

Institutional Review Board of Northwestern University gave ethical approval for this work.

